# Association of radiologic findings with mortality of patients infected with 2019 novel coronavirus in Wuhan, China

**DOI:** 10.1101/2020.02.22.20024927

**Authors:** Mingli Yuan, Wen Yin, Zhaowu Tao, Weijun Tan, Yi Hu

## Abstract

**Background:** Radiologic characteristics of 2019 novel coronavirus (2019-nCoV) infected pneumonia (NCIP) which had not been fully understood are especially important for diagnosing and predicting prognosis.

**Purpose:** To summarize the clinical and radiologic characteristics of NCIP cases and analyze the association of radiologic findings with mortality cases.

**Material and methods:** We retrospective studied 27 consecutive patients who were confirmed NCIP, the clinical characteristics and CT image findings were collected, and the association of radiologic findings with mortality of patients was evaluated.

**Results:** 27 patients included 12 men and 15 women, with median age of 60 years (IQR 47-69). 17 patients discharged in recovered condition and 10 patients died in hospital. The median age of mortality group was higher compared to survival group (68 (IQR 63-73) vs 55 (IQR 35-60), P = 0.003). The comorbidity rate in mortality group was significantly higher than in survival group (80% vs 29%, P = 0.018). The predominant CT characteristics consisted of ground glass opacity (67%), bilateral sides involved (86%), both peripheral and central distribution (74%), and lower zone involvement (96%). The median CT score of mortality group was higher compared to survival group (30 (IQR 7-13) vs 12 (IQR 11-43), P = 0.021), with more frequency of consolidation (40% vs 6%, P = 0.047) and air bronchogram (60% vs 12%, P = 0.025). An optimal cutoff value of a CT score of 24.5 had a sensitivity of 85.6% and a specificity of 84.5% for the prediction of mortality.

**Conclusions:** 2019-nCoV was more likely to infect elderly people with chronic comorbidities. CT findings of NCIP were featured by predominant ground glass opacities mixed with consolidations, mainly peripheral or combined peripheral and central distributions, bilateral and lower lung zones being mostly involved. A simple CT scoring method was capable to predict mortality.

## Introduction

In December, 2019, a series of pneumonia cases linked to a seafood and wet animal wholesale market emerged in Wuhan, Hubei, China ^[1]^. Deep sequencing analysis from lower respiratory tract samples indicated a novel coronavirus, which was named 2019 novel coronavirus (2019-nCoV) ^[2]^. By 4 Feb 2020 a total 24,324 cases have been confirmed infection nationwide with 3,219 severe cases, 490 death cases, and another 23,260 suspected cases ^[3]^. Reports have also been released of exported cases in many other countries. WHO had determined this situation should be deemed a public health emergency of international concern on 30 Jan 2020 ^[4]^. Symptoms of 2019-nCoV infected pneumonia (NCIP) include fever and/or respiratory illness, lymphopenia, and radiologic abnormality ^[5-7]^, among these clinical features, radiologic characteristics of NCIP are especially important for diagnosing and predicting prognosis. We herein summarize the clinical and radiologic characteristics of 27 confirmed cases and analyze the association of radiologic findings with mortality cases.

## Methods

### Patients

This retrospective study was approved by the Institutional Review Boards of Hubei Public Health Clinical Center, the central Hospital of Wuhan, and written informed consent was waived. Between 1 January 2020 and 25 January 2010, 27 consecutive patients who fulfilled the clinical criteria for 2019 novel coronavirus (2019-nCoV) infected pneumonia (NCIP) established by the WHO interim guidance ^[8]^ and discharged with recovered symptoms or died in hospital were included. Diagnosis of NCIP was made by positive test for 2019-nCoV viral RNA, in throat-swab specimens collected from patients by real-time reverse transcription polymerase chain reaction (RT-PCR) using standard RTPCR protocol at Hubei Provincial Center for Disease Control and Prevention (CDC). Clinical characteristics together with chest imaging manifestations of each confirmed cases were recorded, if available.

All patients were treated with intravenous ribavirin 0.5g twice daily and/or oral oseltamivir 75 mg, twice daily. Antibiotics, including levofloxacin, moxifloxacin, sulbactam and cefoperazone, piperacillin and meropenem were used. Some patients also received glucocorticoid and/or intravenous immunoglobulin administration.

### CT scans

The technical parameters included 64-section scanner with 1 mm collimation at 5 mm intervals. All CT examinations were performed with the patient in the supine position and with breath-holding following inspiration, without administration of contrast material. Images were obtained with both mediastinal (width 350 HU; level 40 HU) and parenchymal (width 1500HU; level −600 HU) window settings.

### Imaging evaluation

Two experienced pulmonologists (YML, WY) reviewed the images independently, with a final finding reached by consensus when there was a discrepancy. CT findings included ground glass opacity, consolidation, air bronchogram and nodular opacities. Ground glass opacity (GGO) was defined as hazy areas of increased opacity or attenuation without concealing the underlying vessels. Consolidation was defined as homogeneous opacification of the parenchyma obscuring the underlying vessels. Air bronchogram was defined as a pattern of air-filled (low-attenuation) bronchi on a background of opaque (high-attenuation) airless lung. Nodular opacities were defined as focal round opacities either solid nodules or GGOs (diameter ≤ 3 cm). The presence of lymphadenopathy was defined as a lymph node ≥1 cm in short-axis diameter.

The CT scans were scored on the axial images referring to the method described previously ^[9]^. The extent of involvement of each abnormality was assessed independently for each of 3 zones: upper (above the carina), middle (below the carina and above the inferior pulmonary vein), and lower (below the inferior pulmonary vein). The location of the lesion was defined as peripheral if it was in the outer one-third of the lung, or as central otherwise. The CT findings were graded on a 3-point scale: 1 as normal attenuation, 2 as ground-glass attenuation, and 3 as consolidation. Each lung zone, with a total of six lung zones in each patient, was assigned a following scale according to distribution of the affected lung parenchyma: 0 as normal, 1 as < 25% abnormality, 2 as 25–50% abnormality, 3 as 50–75% abnormality, and 4 as > 75% abnormality. The four-point scale of the lung parenchyma distribution was then multiplied by the radiologic scale described above. Points from all zones were added for a final total cumulative score, with value ranging from 0 to72 (Figure 1).

**Figure 1.**
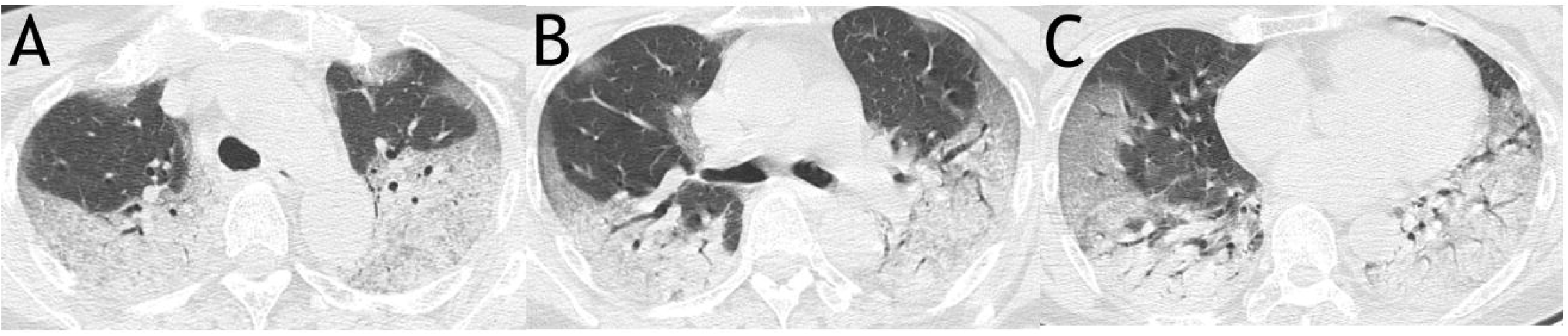
A sample scoring on CT images of a 63-year-old woman from mortality group demonstrated a total score of 63. It was calculated as: for upper zone (A), 3 (consolidation) × 3 (50-75% distribution) × 2 (both right and left lungs) + 2 (ground glass opacity) ×1 (< 25% distribution) × 2 (both right and left lungs); for middle zone (B), 3 (consolidation) × 2 (25-50% distribution) × 2 (both right and left lungs) + 2 (ground glass opacity) × 2 (25-50% distribution) × 2 (both right and left lungs); for lower zone (C), 3 (consolidation) × (2 (25-50% distribution of the right lung) + 3 (50-75% distribution of the left lung)) + 2 (ground glass opacity) × (2 (25-50% distribution of the right lung) + 1 (< 25% distribution of the left lung)).

### Statistical analysis

Continuous variables were expressed as median (interquartile range, IQR) and compared with the Mann-Whitney U test or Wilcoxon test; categorical variables were expressed as number (%) and compared by χ^2^ test or Fisher’s exact test if appropriate. A two-sided α of less than 0.05 was considered statistically significant. Statistical analyses were done using the SPSS (version 16.0).

## Results

As shown in Table 1, the 27 patients diagnosed as 2019 novel coronavirus (2019-nCoV) infected pneumonia (NCIP) included 12 men and 15 women, with median age of 60 years (IQR 47-69). 17 patients discharged in recovered condition with twice negative test for 2019-nCoV viral RNA in throat-swab specimens and 10 patients died in hospital. The median age of mortality group was higher compared to survival group (68 (IQR 63-73) vs 55 (IQR 35-60), P = 0.003). The comorbidity rate in mortality group was significantly higher than in survival group (80% vs 29%, P = 0.018), especially comorbid hypertension, diabetes, and cardiac disease. 12 patients developed acute respiratory distress syndrome and required non-invasive ventilator mechanical ventilation, 11 of whom died, and one 36 years old man (oxygenation index: 184 mmHg) recovered after treatment. There were no significant differences between survival group and mortality group with respect to patient sex and symptoms.

**Table 1.** Demographics, comorbidities, and clinical presentations in mortality and survival groups.

| Variable | All patients<br>N=27 | Survival group<br>N=17 | Mortality group<br>N=10 | P value |
| --- | --- | --- | --- | --- |
| Age, median(IQR), years | 60 (47-69) | 55 (35-60) | 68 (63-73) | 0.003 |
| Male sex | 12 (45) | 8 (47) | 4 (40) | 1.000 |
| Comorbidity | 13 (48) | 5 (29) | 8 (80) | 0.018 |
| Hypertension | 5 (19) | 0 | 5 (50) | 0.003 |
| Diabetes | 6 (22) | 0 | 6 (60) | 0.001 |
| Cardiac disease | 3 (11) | 0 | 3 (30) | 0.000 |
| Tumor | 1 (4) | 1 (6) | 0 | 1.000 |
| Cerebral infarction | 1 (4) | 0 | 1 (10) | 0.370 |
| Chronic gastritis | 1 (4) | 1 (6) | 0 | 1.000 |
| ARDS | 11 (41) | 1 (6) | 10 (100) | 0.000 |
| Symptoms |  |  |  |  |
| Fever | 21 (78) | 15 (88) | 6 (60) | 0.153 |
| Cough | 16 (59) | 11 (65) | 5 (50) | 0.687 |
| Myalgia | 3 (11) | 2 (12) | 1 (10) | 1.000 |
| Dyspnea | 11 (41) | 1 (6) | 10 (100) | 1.000 |

Percentages are in parentheses, unless indicated as median (interquartile range, IQR).
ARDS: acute respiratory distress syndrome.

The CT features, including the location, extent and distribution of involvement of each abnormality were shown in table 2. The median days of CT scans were 8 (IQR 5-11) before symptom onset in all patients. The predominant CT characteristics consisted of ground glass opacity (67%), bilateral sides involved (86%), both peripheral and central distribution (74%), and lower zone involved (96%). Ground glass nodular opacities were found in a 2 cases on admission in survival group and progressed to multiple patchy ground glass opacities on reexamination (Figure 2). Lymphadenopathy and pleural effusion were relatively seldom seen (0% and 4%). The median CT score of mortality group was higher compared to survival group (30 (IQR 7-13) vs 12 (IQR 11-43), P = 0.021), with more frequency of consolidation (40% vs 6%, P = 0.047) and air bronchogram (60% vs 12%, P = 0.025) (Figure 3).

**Table 2.** CT features in mortality and survival groups.

| CT features | All patients<br>N = 27 | Survival group<br>N = 17 | Mortality group<br>N = 10 | P Value |
| --- | --- | --- | --- | --- |
| Symptom onset before CT,<br>median (IQR), days | 8 (5-11) | 9 (5.5-10.5) | 7.5 (3-15) | 0.919 |
| CT score, median (IQR) | 12 (8-29) | 12 (7-13) | 30 (11-43) | 0.021 |
| GGO | 18 (67) | 12 (71) | 6 (60) | 0.683 |
| Consolidation | 5 (19) | 1 (6) | 4 (40) | 0.047 |
| both | 8 (30) | 4 (24) | 4 (40) | 0.415 |
| Air bronchogram | 8 (30) | 2 (12) | 6 (60) | 0.025 |
| Nodular opacities | 2 (7) | 2 (12) | 0 | 0.516 |
| Lymphadenopathy | 0 | 0 | 0 |  |
| Pleural effusions | 1 (4) | 1 (6) | 0 | 1.000 |
| Unilateral | 0 | 0 | 0 |  |
| Bilateral | 1 (4) | 1 (6) | 0 | 1.000 |
| Anatomic sides involved |  |  |  |  |
| Unilateral | 4 (15) | 4 (24) | 0 | 0.264 |
| Bilateral | 23 (86) | 13 (76) | 10 (100) | 0.264 |
| Predominant distribution |  |  |  |  |
| Central | 0 | 0 | 0 |  |
| Peripheral | 7 (26) | 6 (35) | 1 (10) | 0.204 |
| Both | 20 (74) | 11 (65) | 9 (90) | 0.204 |
| Involved zone |  |  |  |  |
| Upper | 22 (81) | 14 (82) | 8 (80) | 1.000 |
| Middle | 23 (85) | 15 (88) | 9 (90) | 1.000 |
| Lower | 26 (96) | 16 (94) | 10 (100) | 1.000 |

Percentages are in parentheses, unless indicated as median (interquartile range, IQR).
GGO: ground glass opacity.

**Figure 2.**
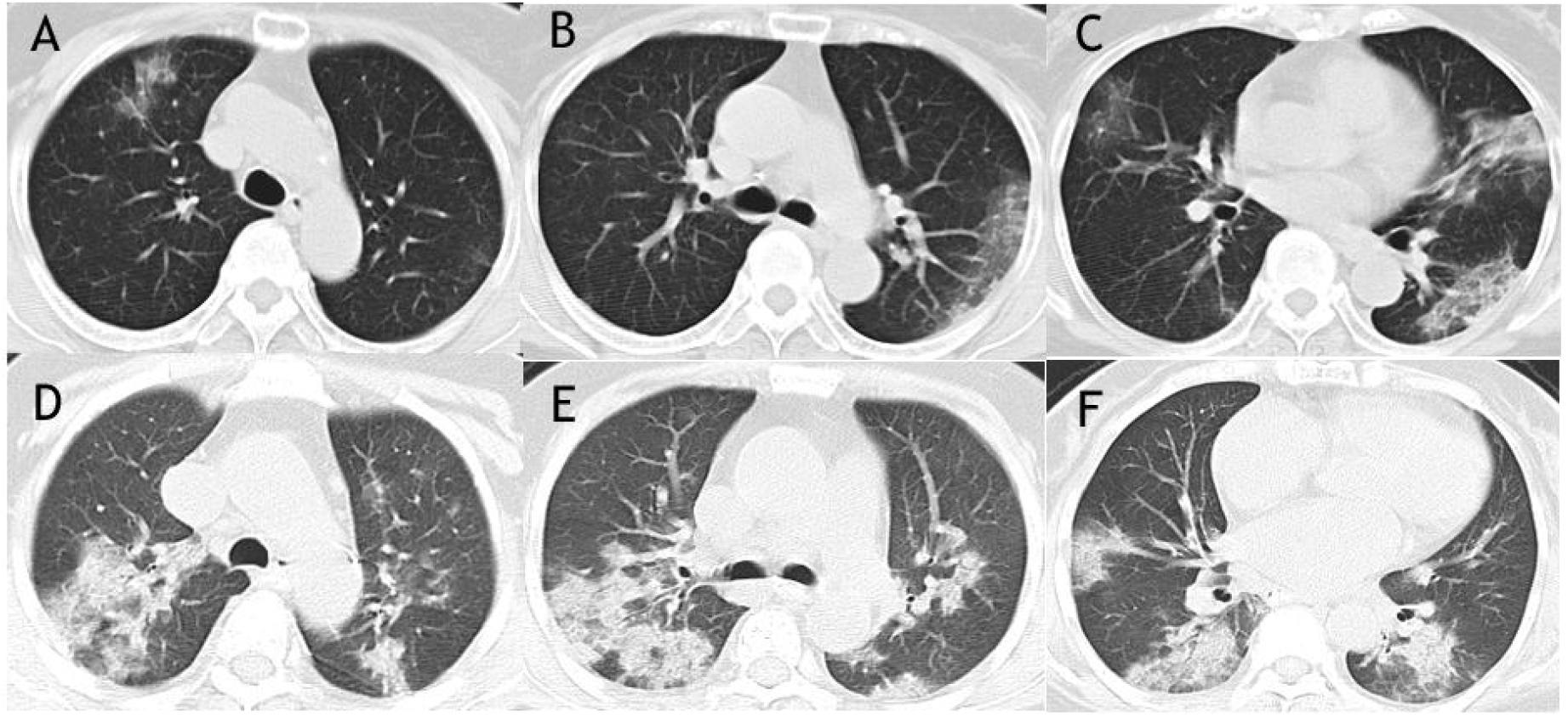
Two cases of CT images from survival group showed ground glass nodular opacities on admission and progressed to multiple patchy ground glass opacities on reexamination. The CT images of a 60-year-old woman on admission showed peripherally distributed focal ground glass nodular opacities in only right lung (A) and rechecked CT images showed expanded area of patchy ground glass opacities in both right and left lungs with reticular and interlobular septal thickening on 7 days later (B). Unilateral ground glass nodular opacity was found on CT images of a 64-year-old man from survival group (C) and progressed patchy ground glass opacities as well as interlobular septal thickening were seen after 4 days (D).

**Figure 3.**
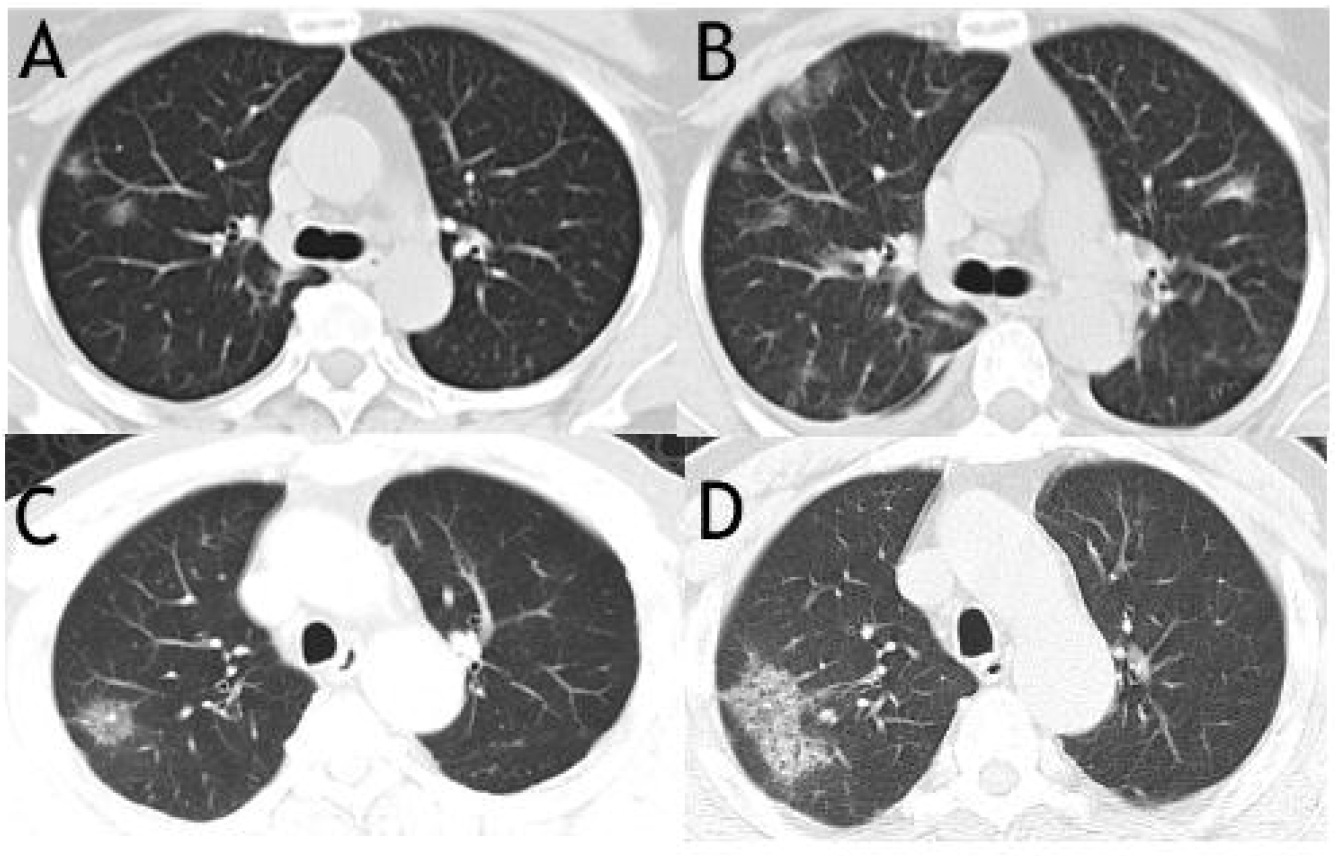
CT images of a 76-year-old woman from survival group showed pure ground glass opacities with predominant peripheral distribution in middle and lower lung zones (A-C). Air bronchogram, together with extensive of consolidations and ground glass opacities were found in the CT images of a 72–year-old woman from mortality group (D-F).

In the receiver operating characteristic curve analysis (Figure 4), an optimal cutoff value of a CT score of 24.5 had a sensitivity of 85.6% and a specificity of 84.5% for the prediction of mortality. The area under the receiver operating characteristic curve was 0.901 (95% confidence interval: 0.873, 0.928).

**Figure 4.**
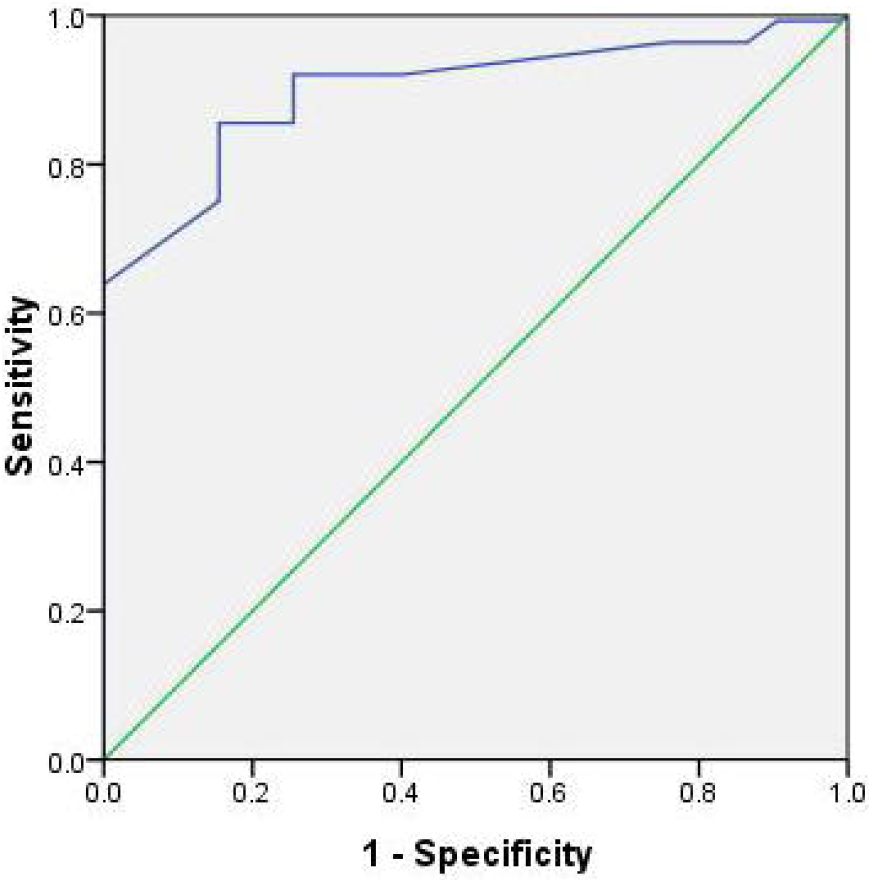
ROC analysis of the CT score for prediction of mortality. N = 27; AUC=0.901.

Initial and reexamined CT features in survival group were compared in Table 3. 11 patients reexamined CT scans on day 15 (IQR 9-18) after symptom onset, the morphology of the lesions, the location, extent and distribution of involvement of each abnormality were not significantly changed compared to those on admission. In mortality group, CT scores progressed rapidly in a short time (12 (IQR 5-24.5) vs 20 (IQR 15-46), P = 0.042), with more lung zones being involved (Table 4).

**Table 3.** Comparison of CT features in survival group between admission and reexamination.

| CT features | Admission<br>N = 11 | Reexamination<br>N = 11 | P Value |
| --- | --- | --- | --- |
| Symptom onset before CT, median (IQR), days | 7 (5-10) | 15 (9-18) |  |
| CT score, median (IQR) | 12 (6-12) | 12 (6-15) | 0.237 |
| GGO | 8 (73) | 7(64) | 1.000 |
| Consolidation | 1(9) | 0 | 1.000 |
| both | 2(18) | 4 (36) | 0.635 |
| Air bronchogram | 1(9) | 0 | 1.000 |
| Nodular opacities | 2 (18) | 1 (9) | 1.000 |
| Lymphadenopathy | 0 | 0 |  |
| Pleural effusions | 0 | 1 (9) | 1.000 |
| Unilateral | 0 | 0 |  |
| Bilateral | 0 | 1 (9) | 1.000 |
| Anatomic sides involved |  |  |  |
| Unilateral | 3 (27) | 2 (18) | 1.000 |
| Bilateral | 8(73) | 9 (82) | 1.000 |
| Predominant distribution |  |  |  |
| Central | 0 | 0 |  |
| Peripheral | 5 (45) | 4 (36) | 1.000 |
| Both | 6 (55) | 7(64) | 1.000 |
| Involved zone |  |  |  |
| Upper | 7(64) | 7(64) | 1.000 |
| Middle | 8(73) | 11 (100) | 0.214 |
| Lower | 10(91) | 10 (91) | 1.000 |

Percentages are in parentheses, unless indicated as median (interquartile range, IQR).
GGO: ground glass opacity.

**Table 4.** Comparison of CT features in mortality group between admission and reexamination.

| CT features | Admission<br>N = 5 | Reexamination<br>N = 5 | P Value |
| --- | --- | --- | --- |
| Symptom onset before CT, median (IQR), days | 8 (2-22.5) | 11 (4.5-27.5) |  |
| CT score, median (IQR) | 12 (5-24.5) | 20 (15-46) | 0.042 |
| GGO | 4 (80) | 4 (80) | 1.000 |
| Consolidation | 0 | 0 |  |
| both | 1 (20) | 1 (20) | 1.000 |
| Air bronchogram | 1 (20) | 1 (20) | 1.000 |
| Nodular opacities | 0 | 0 |  |
| Lymphadenopathy | 0 | 0 |  |
| Pleural effusions | 0 | 0 |  |
| Unilateral | 0 | 0 |  |
| Bilateral | 0 | 0 |  |
| Anatomic sides involved |  |  |  |
| Unilateral | 0 | 0 |  |
| Bilateral | 5 (100) | 5 (100) | 1.000 |
| Predominant distribution |  |  |  |
| Central | 0 | 0 |  |
| Peripheral | 1 (20) | 0 | 1.000 |
| Both | 4 (80) | 5 (100) | 1.000 |
| Involved zone |  |  |  |
| Upper | 3 (60) | 5 (100) | 0.444 |
| Middle | 4 (80) | 5 (100) | 1.000 |
| Lower | 5 (100) | 5 (100) | 1.000 |

Percentages are in parentheses, unless indicated as median (interquartile range, IQR).
GGO: ground glass opacity.

## Discussion

Coronaviruses (CoV) are a large family of viruses that cause illness ranging from the common cold to more severe diseases such as Middle East Respiratory Syndrome (MERS-CoV) and Severe Acute Respiratory Syndrome (SARS-CoV). A novel coronavirus (nCoV) is a new strain that has not been previously identified in humans ^[10]^. The 2019-nCoV causes symptoms similar to SARS based on recent clinical data ^[5, 6]^, and is capable of spreading from human to human and between cities, with very contagious characteristic ^[11]^. The mortality of the 27 included patients infected by 2019-nCoV was 37%, which is much higher than that reported 2% on 4 Feb 2020 ^[3]^. Giving the priority of 2019-nCoV detection to severe cases because of shortness of test kits in our hospital initially and potential false negative results of 2019-nCoV viral RNA detection in mild cases might account for this discrepancy. Consistent with recent reports, our results suggest that 2019-nCoV is more likely to infect elderly people with chronic comorbidities as a result of the weaker immune functions of these patients ^[5, 6]^, and 2019-nCoV-associated death is also related to elder age and underlying illnesses, especially hypertension, diabetes, and cardiac disease.

Consistently with several recent reports regarding to CT findings of 2019-nCoV infected pneumonia (NCIP) ^[12-14]^, our results showed that CT manifestations of NCIP were featured by predominant ground glass opacities (GGO) (67%) mixed with consolidations (30%), mainly peripheral (26%) or combined peripheral and central distributions (74%), bilateral (86%) and lower lung zones (96%) being mostly involved. Merely consolidation, central distribution only, pleural effusions or lymphadenopathy were relatively rarely seen. Initially the CT images might be GGO nodules or patchy GGO mostly peripherally distributed then consolidations and extensive distributions were seen when pneumonia progressed. Consolidation, air bronchogram, extensive distribution as well as multiple involved lung zones were more common in mortality group, suggesting a more severe clinical course for these abnormalities can be pathologically correlated with diffuse alveolar damage and similar conclusions can be drawn in H1N1 pneumonia, N5N1 pneumonia, H7N9 pneumonia and SARS ^[9, 15-17]^. As members of the same virus family, NCIP shared much common ground with MERS and SARS in CT features. Subpleural and basilar airspace lesions with GGOs and consolidations were conspicuous CT characteristics of all three kinds of virus pneumonia, but pleural effusions are more common in patients who died of MERS ^[18-21]^.

Comprehensively evaluated the CT features of NCIP, we calculated the CT score of each patient. The CT scores were much higher in mortality group compared to survival group (30 (IQR 7-13) vs 12 (IQR 11-43), 0.021), P = 0.021) on admission. In mortality group the scores markedly increased in a short time (12 (IQR 5-24.5) vs 20 (IQR 15-46), P = 0.042), suggesting a progressive course of NCIP. Although the CT scores were not diminished, they maintained a relatively stable level for a period of time in survival group (12 (IQR 6-12) vs 12 (IQR 6-15), P = 0.237), suggesting that it might take a long time to recover in chest imaging when NCIP was under controlled. We found an optimal cutoff value of a CT score of 24.5 (sensitivity of 85.6% and specificity of 84.5%) to predict mortality. We hope the simple scoring method according to CT scans may help triage patients and screening patients who need more aggressive treatment and closely monitoring. However, the efficacy of such approach to decrease mortality remains to be validated in future studies.

Although a definite diagnosis of NCIP cannot be achieved by using imaging features alone for most viral pneumonia imaging patterns share similarity, the CT features summarized above might be helpful in differentiation various pathogens of pneumonia and in triage patients.

There were limitations in our study. First it was a retrospective study including only 27 inpatients, while outpatients and suspected but undiagnosed cases for deficiency of detection kits of 2019-nCoV and potential false negative results were ruled out. Second, not all information was collected because a significant part of patients had not rechecked CT scans. Then, it was pulmonologists, although they are experienced, who evaluated CT images. Finally there was a lack of information regarding interobserver agreement, because the study emphasis on the final consensus interpretation rather than independent reading.

## Data Availability

Partial data of this study are included within the article and all data can be supplied if requested.

